# Enhancing Polygenic Risk Prediction by Modeling Quantile-Specific Genetic Effects

**DOI:** 10.64898/2025.12.25.25342935

**Authors:** Suin Kim, Taewan Goo, Taesung Park, Mira Park

## Abstract

Polygenic risk scores (PRSs) quantify an individual’s genetic susceptibility to complex traits and diseases. Conventional PRSs, which are based on linear models, perform poorly for phenotypes with skewed distributions or with genetic effects that vary across the distribution. We propose a quantile regression-based PRS (QPRS) that can capture quantile-specific genetic effects. While existing PRSs provide only a single score, QPRS models genetic influences at multiple quantiles of the phenotype, thereby enhancing predictive performance by utilizing these multiple scores as covariates. We evaluate the performance of our method through both simulations and a real-data application. In simulations, QPRS significantly reduces the mean squared error (MSE) compared to the conventional PRS, both in the presence of variance quantitative trait loci and outliers. For real data analysis, we use data from Korea Genome and Epidemiology Study (KoGES) to evaluate predictive performance. We consider two prediction tasks: a continuous outcome (glucose level) and a binary outcome (diabetes status, derived from glucose level). For glucose-level prediction, the model incorporating QPRS achieves a *R*^2^ value 4.69 times higher than the model using conventional PRSs. For predicting diabetes status, the model with QPRS produces an area under the curve 1.06 times higher than the model with conventional PRSs.

## 1. Introduction

Polygenic risk scores (PRSs) are a widely used tool for translating genome-wide association study (GWAS) findings into individual-level predictions of complex traits and disease risk. By aggregating genome-wide variant effects, particularly those of single-nucleotide polymorphisms (SNPs), PRSs can help identify individuals at elevated risk, guide preventive strategies, and contribute to personalized medicine^1–3^. A variety of statistical approaches exist for PRS construction, including clumping-and-thresholding, lassosum^4^, LDpred^5^, and SBLUP^6^. These methods differ in how they model linkage disequilibrium (LD), apply shrinkage priors, and implement penalized regression. These methods have been applied in various contexts, including predicting body mass index and obesity outcomes using lassosum^7^, analyzing a range of complex traits in the UK Biobank^8^, and assessing the transferability of PRS estimates for height and BMI across populations^9^.

Despite these advances, most PRS construction methods rely on GWAS based on ordinary least squares regression or related linear models, which focus on conditional mean effects^10^. This mean-centric perspective can mask heterogeneity when genetic effects vary across the phenotype distribution. For example, some variants may exert a stronger influence on the distribution tails rather than the center, yet such distributional differences are overlooked in mean-based models. Several studies have explored heterogeneity in genetic effects across trait distributions, reporting evidence that genetic influences may differ substantially depending on the quantile of the phenotype considered^11–14^.

Quantile regression (QR)^15,16^ offers a complementary perspective by estimating the genetic effects across conditional quantiles of the phenotype distribution. This approach provides a distribution-aware view of genetic associations that conventional mean-based models may not capture. QR has been used in estimating generic effect to flowering-time traits in common bean^17^, to body mass index^11^, and 28 continuous quantitative traits including blood glucose, total cholesterol, BMI^18^. Recently, Wang, et al. ^19^ proposed QR-based GWAS summary statistics derived from the UK Biobank data. However, these works primarily address discovery rather than prediction. To our knowledge, no cohesive predictive framework exists that directly integrates quantile-specific genetic effects into a PRS through the aggregation of information across multiple quantiles. Mefford, et al. ^20^ applied conventional PRS in a quantile-prediction setting; however, the PRS weights were derived from mean-regression GWAS, and thus their model did not incorporate quantile-specific effects of SNPs.

We propose a quantile regression-based PRS (QPRS) that incorporates quantile-specific genetic effects identified from quantile regression-based GWAS (QR-GWAS) into the risk score’s construction. We construct risk scores from QR-GWAS summary statistics using a clumping-and-thresholding (C+T) procedure, the most commonly used method for computing polygenic scores. By incorporating risk scores from multiple quantiles as covariates in a phenotype prediction, QPRS captures distributional heterogeneity and efficiently pools information, thereby extending conventional PRS beyond mean-based associations. We evaluate its performance in two settings: (i) simulation studies comparing QPRS with conventional PRSs, and (ii) a real-data application using genome-wide data from the Korean Association Resource (KARE) cohort of the Korean Genome and Epidemiology Study (KoGES).

## 2. Materials and Methods

### 2.1 Data Description

We use genotype data from the KARE cohort, a population-based study within the KoGES investigating genetic determinants of complex traits and diseases. The data are accessed through the Clinical and Omics Data Archive (CODA; https://coda.nih.go.kr) of the Korea Disease Control and Prevention Agency (KDCA).

The dataset originally consisted of 8,840 participants and 1,573,861 SNPs. Genotypes are imputed with the Asian 1000 Genomes reference panel^21^. We apply standard quality control (QC) procedures: we exclude individuals with ≥10 missing genotypes or incomplete covariate information, and filter variants with a minor allele frequency (MAF) <1, a Hardy-Weinberg equilibrium exact test p-value < 1 × 10^−6^, or missing data >5. In addition, individuals taking oral diabetes medication are excluded. After QC, 8,408 individuals and 1,573,859 SNPs remain. For subsequent analyses, including both the simulation study and real-data evaluation, we randomly divide the 8,408 individuals into a discovery set (6,000 individuals) and a target set (2,408 individuals) in each iteration. In the real-data analysis, these iterations correspond to the repetitions within 5-fold cross-validation, while in the simulation study, they represented independent simulation replicates.

### 2.2 Quantile-based Polygenic Risk Score (QPRS)

PRSs are typically defined as a weighted sum of risk alleles, where the weights are SNP effect sizes derived from GWAS summary statistics^22^. PRS construction typically involves two data sources: a discovery set, from which summary statistics are derived, and a target set, in which the PRS is calculated for each individual and subsequently evaluated^23^.

The proposed QPRS extends this standard framework by incorporating quantile-specific effect sizes, thereby capturing heterogeneous genetic effects across the different parts of the phenotype distribution. The construction procedure of QPRS consists of three steps: (i) obtaining QR-GWAS summary statistics, (ii) computing QPRS with selected variants, and (iii) using these scores as joint covariates in a downstream prediction model. The key point is to use scores derived at multiple quantile levels to capture distributional heterogeneity and improve predictive performance. Figure 1 illustrates the overall workflow of QPRS construction, from GWAS preprocessing to quantile-specific score calculation.

**Figure 1.**
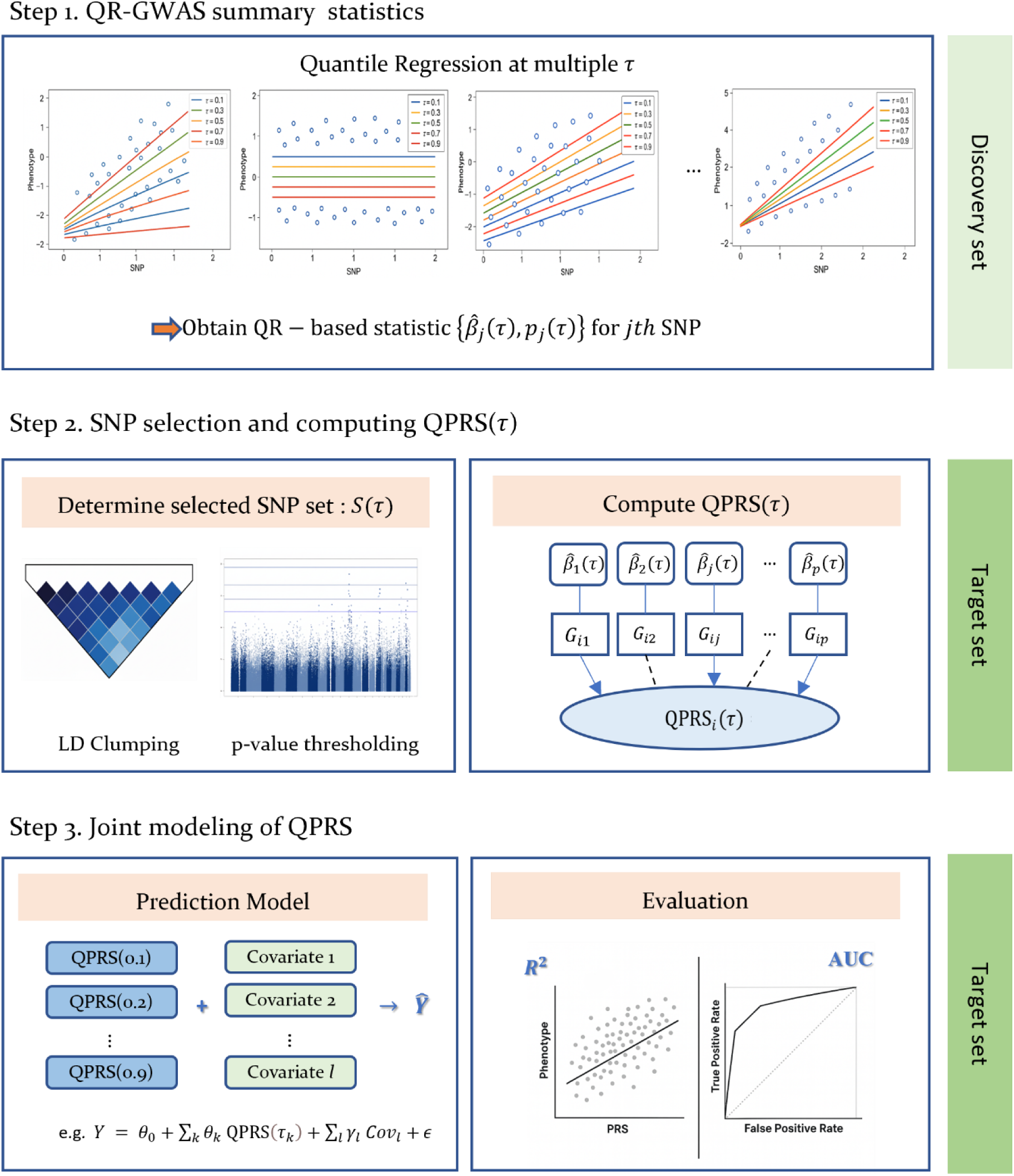
Workflow of QPRS construction. The QPRS estimates SNP effects at multiple quantiles, performs C+T to select variants, and combines quantile-wise PRSs for prediction in the target set.

#### Step 1: QR-GWAS Summary Statistics

In the first step, we conduct genome-wide QR on the discovery set at *τ* = 0.1, …,0.9 to obtain GWAS summary statistics. Let *Y*_*i*_ denote the phenotype, *G*_*ij*_ genotype dosage for SNP *j* (coded as 0, 1, or 2 effect allele), and *C*_*i*_ covariates (e.g., age, sex, or principal components of ancestry). The *τ* -th conditional quantile function is modeled as

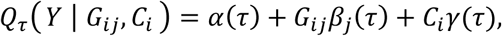

where *⍺*(*τ*) is the intercept, *β*_*j*_(*τ*) the SNP effect at quantile *τ*, and *γ*(*τ*) the covariate effects at quantile *τ*. The regression coefficients are estimated by solving the optimization problem

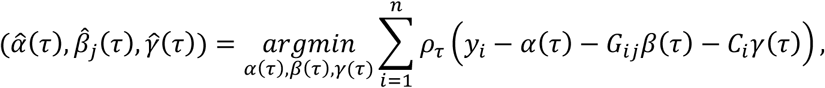

where *ρ*_*τ*_(*u*) = *u*(*τ* − **1**_{*u*<0}_) is the check loss, the standard loss function in QR. Here, **1**_{*u*<0}_is the indicator function, which equals 1 if *u* < 0 and 0 otherwise. The loss function weights positive and negative residuals asymmetrically, allowing estimation of conditional quantiles rather than the conditional mean. Statistical significance of *β̂*_*j*_(*τ*) is evaluated using the rank-score test^24^, yielding a *p*-value *p*_*j*_(*τ*) for SNP *j*. Accordingly, QR-GWAS produces summary statistics {*β̂*_*j*_(*τ*), *p*_*j*_(*τ*): *j* = 1, …, *m*}, where *m* is the total number of SNPs.

In addition to the approach adopted here, several alternative methods are available. For instance, QR-GWAS summary statistics can be obtained from large-scale published resources^19^, when available, and *p*-values can be derived using Wald-type tests^25^, which are based on asymptotic standard errors.

#### Step 2: Computing QPRS and SNP Selection

We construct quantile-specific risk scores at *τ* = 0.1, …, 0.9. For a given quantile *τ* ∈ (0,1), the QPRS for individual *i* is defined as

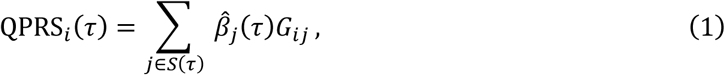

where *G*_*ij*_ is the genotype dosage of SNP *j* for individual *i*, *S*(*τ*) is the set of selected SNPs at quantile *τ* after the selection procedure, and *β̂*_*j*_(*τ*) is the quantile-specific effect size of the *j*-th SNP obtained from the QR-GWAS summary statistics in step 1. This formulation extends the conventional PRS by allowing genetic effects to vary across the phenotype quantiles. For notational simplicity, we suppress the individual index *i* hereafter.

To obtain *S*(*τ*), we employ a C+T strategy, a widely used and computationally straightforward method for PRS construction^26^. First, SNPs in high LD are clumped by retaining the most significant variant within each LD block. Next, variants are filtered based on a predefined *p*-value threshold. Within each LD block, we select the SNP that has the smallest *p*-value and is below the threshold. A common threshold can be imposed across all quantiles, as in this study, or specific thresholds may be applied to each quantile. In line with standard C+T practice, we consider multiple thresholds and retain the one that yields the best predictive performance.

At each quantile, the C+T procedure is repeated to identify informative variants, yielding a quantile-specific SNP set *S*(*τ*). Because effect sizes and corresponding *p*-values differ across quantiles, the resulting SNP sets vary accordingly. The C+T procedure can be implemented with standard tools such as PLINK, since effect-size estimates and *p*-values were already obtained in step 1. Although these estimates are derived from QR, they are provided in a format directly comparable to those from conventional linear regression–based GWAS. In this study, we apply the C+T approach to keep the procedure simple and to focus on presenting the methodology. More advanced selection methods are possible and will be discussed in Section 4.

#### Step 3: Joint modeling of QPRS

Each QPRS(*τ*) represents the genetic contribution to a specific conditional quantile of the phenotype, thereby offering insights beyond the mean effect captured by conventional linear PRS. While individual quantile-specific scores provide valuable distributional information, their utility is further enhanced when considered together. We therefore adopt a joint modeling framework that simultaneously incorporates QPRS across multiple quantiles. For *τ*_*k*_ = *k*/10, the phenotype is predicted as

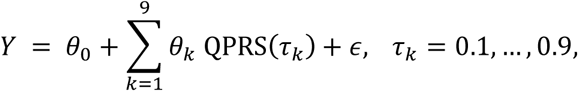

where *θ*_0_ is an intercept and *θ*_*k*_ is a regression coefficient for QPRS(*τ*_*k*_). This formulation integrates information across quantiles, improving prediction and robustness while preserving the interpretability of each quantile-specific effect. Unless otherwise specified, “QPRS” hereafter refers to this joint modeling framework. Additional covariates, such as sex and age, can also be incorporated into the model. The regression framework is selected based on the outcome type: linear regression for continuous traits, logistic regression for binary traits, and QR for predicting phenotype quantiles.

The key distinction between QPRS and conventional PRS is that multiple genetic scores can be incorporated as covariates in the predictive model. Unlike the conventional approach representing genetic effects only on single mean-based score, QPRS employs multiple quantile-based scores. Thus, QPRS captures heterogeneity in genetic effects enabling effective application to various prediction tasks, such as predicting conditional quantiles of the phenotype or its mean. Each QPRS(*τ*) reflects genetic effects on a different quantile of the phenotype, and together they enhance prediction accuracy. As shown in Section 3, this joint modeling improves predictive accuracy.

### 2.3 Simulation study

We evaluate the proposed QPRS method using Monte Carlo simulations. We also consider a conventional PRS from mean regression using the C+T procedure, hereafter denoted as the linear regression-based PRS (LPRS).

To mimic realistic genetic architecture, we generate semi-synthetic datasets by sampling genotypes from the KARE dataset while retaining key features such as the minor allele frequency spectrum. A comprehensive description of the KARE dataset is provided in Section 2.1. For the simulation study, we reduce the dimensionality of the genotype data by applying LD pruning. This step mitigates spurious correlations due to LD and preserves causal SNPs, enabling a more reliable assessment of whether the QR-based C+T procedure could accurately select the causal SNPs. Using a sliding window of 50 SNPs, a step size of 5 SNPs, and an LD threshold of *r*^2^ < 0.2, we retain 154,455 SNPs.

For evaluation, 6,000 individuals are randomly chosen as the discovery set for generating GWAS summary statistics, with the remaining individuals reserved as the target set for PRS calculation and evaluation. We simulate phenotypes under two different schemes. The detailed simulation design and evaluation metrics are provided below.

GWAS summary statistics are obtained separately for each method. For LPRS, we perform GWAS using linear mean regression in the discovery set. For QPRS, we conduct QR-GWAS across multiple quantile levels. In both cases, SNPs are selected using the C+T procedure. LD clumping is performed with a 1 Mb sliding window: within each window, the SNP with the smallest *p*-value is chosen as the index SNP, and all SNPs with pairwise *r*^2^ > 0.5 relative to the index are excluded. This configuration is standard in PRS analyses and corresponds to the default parameter in PLINK^27^.

To account for the sensitivity of C+T to the choice of p-value thresholds, we consider a predefined grid of candidate thresholds. We select different grids of *p*-value thresholds for QPRS and LPRS to account for the distinct distributions of association statistics generated by each method. *P*-values from a standard linear regression-based GWAS (used for LPRS) are not directly comparable to those from the QR-GWAS, which assesses significance at specific quantiles. To ensure a fair comparison, we establish separate but functionally equivalent grids for each method. For LPRS, the grid is: {1 × 10^−6^, 1 × 10^−5^, 5 × 10^−5^, 1 × 10^−4^, 5 × 10^−4^, 1 × 10^−3^, 5 × 10^−3^, 1 × 10^−2^}, while for QPRS, a wider range starting from a more stringent cutoff is used: {1 × 10^−8^, 5 × 10^−8^, 1 × 10^−7^, 1 × 10^−6^, 1 × 10^−5^, 1 × 10^−4^, 1 × 10^−3^, 1 × 10^−2^}. Each grid is designed to span an appropriate spectrum for its respective method, from highly stringent cutoffs yielding few or no SNPs to ones yielding a substantially large number (more than 500 SNPs included). For QPRS, the best-performing threshold is applied uniformly across all quantiles to maintain consistency.

#### Simulation scheme 1: With vQTL

The first simulation setting considers the presence of variance quantitative trait loci (vQTLs), genetic loci that influence the variability of a phenotype rather than its mean. A vQTL effect, by increasing or decreasing phenotypic variance, naturally manifests as heterogeneous genetic effects across the distribution; for instance, a variance-increasing allele will have a larger effect on the upper and lower tails than at the median. This setting allows us to directly assess the ability of QPRS to detect and leverage these heterogeneous effects for improved prediction.

Our simulation design is adapted from Choi and O’Reilly ^28^, with modifications to incorporate heterogeneity. Let *X* denote the genotype matrix. We generate simulated phenotypes as

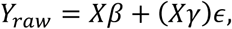

where *β* denotes the mean effect and *γ* the variance effect. Note that when the error term *ε* has mean zero, *γ* does not affect the mean of the phenotype but influences its variance, thereby inducing quantile-specific genetic effects. Specifically, the quantile effect sizes of SNPs are given by

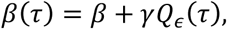

where *Q*_*∈*_(*τ*) is the *τ*-th quantile of the error distribution. For *β*, *p*_1_ causal SNPs are each assigned an effect size of 5/*p*_1_, with all other SNPs set to zero. For *γ*, *p*_2_ causal SNPs are each assigned an effect size of 10/*p*, with the remaining SNPs set to zero. This parameterization ensures the overall magnitude of genetic contribution is comparable across settings, regardless of the number of causal SNPs. By holding the total effect size constant and varying *p*_1_, we contrast sparse architectures, characterized by a few large-effect variants, with polygenic architectures, characterized by many small-effect variants. We fix *p*_2_ = 5 and vary *p*_1_across 5, 10, 50, and 100. For the error term *ε*, we consider three distributions: standard normal, Student’s *t* with 5 degrees of freedom, and standard log-normal.

To adjust for population structure, we regress the simulated phenotype *Y*_*raw*_ on the first five principal components (*PC*_1_,…, *PC*_5_) of the genotype data:

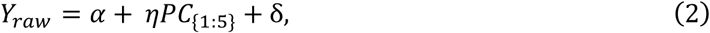

where *⍺* is an intercept, and *η* a coefficient vector of principal components, and δ an error term. Residuals from model (2) are used as the final phenotype *Y*. This adjustment follows the approach of Choi and O’Reilly ^28^.

#### Scheme 2: With Outliers

The second simulation setting considers phenotypes contaminated with massive outliers. Because linear regression is highly sensitive to outliers, it may fail in this scenario, whereas QR provides a more robust alternative and is expected to yield achieve higher predictive accuracy.

We generate phenotypes from the linear model

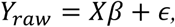

where now *β*(*τ*) = *β* for all *τ* ∈ (0,1), indicating that quantile effects do not vary across quantiles. We assign effect sizes to 50 randomly selected causal SNPs by sampling from the standard normal distribution *N*(0,1), with all other SNP effects set to zero.

To introduce contamination, we multiply phenotype values for 1%, 2%, 5%, or 10% of the observations by factors of 5, 10, or 20. As in equation (2), we use residuals from principal component regression on the first five PCs as the final phenotype.

#### Evaluation metric

We evaluate the performance of the proposed QPRS against the LPRS in terms of SNP selection and predictive accuracy. First, to assess SNP selection performance, we compute the true positive rate (TPR) and false positive rate (FPR). TPR is defined as the proportion of causal SNPs correctly identified after C+T procedure, whereas FPR is the proportion of non-causal SNPs incorrectly selected. Together, these measures quantify how effectively each method distinguishes true genetic signals from background noise.

Next, we evaluate the predictive accuracy of the risk scores. The goal is to determine how effectively the scores predict phenotype outcomes. In scheme 1, we examine the ability of QPRS and LPRS to predict phenotype quantiles at *τ* =0.1 and 0.9. After fitting QR models that include QPRS (or LPRS) along with covariates, predictive accuracy is measured by the mean squared error (MSE) between the estimated and true quantiles. Here, the true quantile function is defined as

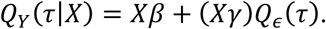

In scheme 2, we assess QPRS and LPRS by their accuracy in predicting the mean and median of the phenotype. After fitting linear regression with QPRS (or LPRS) and covariates, accuracy is quantified by the MSE between the true mean function, *Xβ*, and the fitted values. Each simulation is repeated 10 times, and the reported results are the averages of the evaluation metrics.

### 2.4 Real data analysis

#### Analysis Procedure

We focus on predicting glucose levels two hours after a standard 75g oral glucose tolerance test (OGTT). A 5-fold cross-validation is conducted, stratified by glucose status (>200 mg/dL vs. ≤200 mg/dL). In each fold, four subsets are used as the discovery set for GWAS, adjusting for sex, age, BMI, and the first 10 principal components (*PC*_1_,…, *PC*_10_). The remaining subset serves as the target set, in which we derive and evaluate the PRS.

QPRS is computed following the same procedure as in simulation design. LD clumping is applied in 1 Mb windows, excluding SNPs with *r*^2^ > 0.5 relative to the index SNP. Six *p*-value thresholds for C+T procedure ( 5 × 10^−2^, 1 × 10^−2^, 5 × 10^−3^, 1 × 10^−3^, 1 × 10^−4^, 1 × 10^−5^) are considered. More stringent thresholds are avoided as they yielded almost no variants. The optimal threshold in each fold is selected based on predictive performance, measured by *R*^2^ for the continuous trait and AUC for the binary trait.

#### Evaluation

We evaluate performance for both a continuous trait (2-hour post-OGTT blood glucose level in mg/dL) and a binary trait (defined by binarizing the continuous trait at 200 mg/dL). Clinically, it is a cornerstone for diagnosing type 2 diabetes and prediabetes, with a threshold of 200 mg/dL for defining high-risk status^29^. The distribution of glucose level is often right-skewed, with a tail of high-risk individuals^30^. This characteristic highlights the need for our score, QPRS. By modeling quantile-specific effects, QPRS offers advantages over conventional mean-based methods, which often perform poorly on traits with such skewed distributions.

For the continuous trait, we fit two linear regression models: one including only the PRS as a predictor, and another including the PRS along with covariates (sex, age, BMI, smoking status, and the first 10 principal components, *PC*_1_,…, *PC*_10_). Model performance is assessed using *R*^2^. For the binary trait, we fit logistic regression with the same two configurations (PRS only, and PRS with covariates). For the binary trait, model performance is evaluated using the area under the curve (AUC). We report AUC on two distinct scales to provide a comprehensive assessment. The first is the standard observed-scale AUC, which measures the model’s ability to discriminate between cases and controls within our specific sample. The second is the liability-scale AUC (AUCL)^31^, which is calculated based on the underlying continuous liability threshold model for complex traits. The purpose of using AUCL is to adjust the performance metric for the population prevalence of the disease, thereby mitigating potential biases that can arise from study designs, particularly case-control ascertainment.

#### Comparison Methods

We compare QPRS with four benchmark PRS methods: LPRS, lassosum^4^, LDpred^5^, and SBLUP^6^. All reported metrics are averaged across the five cross-validation folds. We conduct QR analyses using the *quantreg* package^32^ in R, and fit lassosum models with the *lassosum* package^4^ in R version 4.4.0 (R Foundation for Statistical Computing, Vienna, Austria). We perform standard linear regression-based GWAS using PLINK (version 1.90b7.2). For LDpred and SBLUP, we use LDpred^5^ (version 1.0.10) and GCTA^33^, respectively, both of which use GWAS summary statistics generated by BOLT-LMM^34^ as input.

## 3. Results

### 3.1 Simulation Results

#### Scheme 1: With vQTL

Table 1 presents the TPR and FPR of SNPs selected during the C+T procedure of LPRS and QPRS. Across all simulation settings, QPRS achieves consistently higher TPR than LPRS, reflecting an improved ability to identify truly associated variants. This gain in sensitivity persists across different numbers of causal SNPs (*p*_1_) and under various error distributions. In contrast, LPRS often misses a substantial proportion of causal SNPs, especially when errors follow heavy-tailed (*ε* ∼ *t*(5)) or skewed (*∈* ∼ *logN*(0,1)) distributions. The improvement in TPR with QPRS is accompanied by a modest rise in FPR, reflecting the inclusion of some non-causal SNPs. Nevertheless, the gain in TPR generally outweighs this cost, suggesting that QPRS provides a more powerful variant selection strategy than LPRS in the presence of heterogeneous genetic effects.

**Table 1.** True positive rate (TPR) and false positive rate (FPR) of SNP selection for LPRS and QPRS in Scheme 1 across varying numbers of causal SNPs ( *p*_1_) and error distributions. FPR values are multiplied by 10^2^.

| $p_1$ | Method | $\epsilon \sim N(0,1)$ | | $\epsilon \sim t(5)$ | | $\epsilon \sim \log N(0,1)$ | |
| --- | --- | --- | --- | --- | --- | --- | --- |
|  |  | TPR | FPR | TPR | FPR | TPR | FPR |
| 5 | LPRS | 0.380 | 0.004 | 0.300 | 0.002 | 0.160 | 0.005 |
|  | QPRS | 0.850 | 0.019 | 0.790 | 0.015 | 0.840 | 0.029 |
| 10 | LPRS | 0.327 | 0.012 | 0.133 | 0.001 | 0.160 | 0.542 |
|  | QPRS | 0.513 | 0.009 | 0.453 | 0.010 | 0.800 | 1.575 |
| 50 | LPRS | 0.004 | 0.050 | 0.000 | 0.053 | 0.009 | 0.541 |
|  | QPRS | 0.082 | 0.020 | 0.080 | 0.045 | 0.087 | 0.031 |
| 100 | LPRS | 0.001 | 0.001 | 0.003 | 0.103 | 0.003 | 0.070 |
|  | QPRS | 0.035 | 0.014 | 0.037 | 0.016 | 0.045 | 0.068 |

Table 2 presents the predictive accuracy of LPRS and QPRS in the simulation scheme 1. Here, the task is to predict the 0.1 and 0.9 quantiles of the generated phenotype. Across all scenarios, QPRS consistently achieves a lower MSE than LPRS. The improvement is evident regardless of *p*_1_. In other words, QPRS provides stable performance whether the genetic architecture is sparse (a few variants driving the effect) or polygenic (many variants each contributing small effects). The relative advantage of QPRS is especially notable under heavy-tailed and skewed error distributions, where the MSE ratio of LPRS to QPRS exceeds 2 in several cases. LPRS, which assumes normally distributed errors, tends to underperform.

**Table 2.**
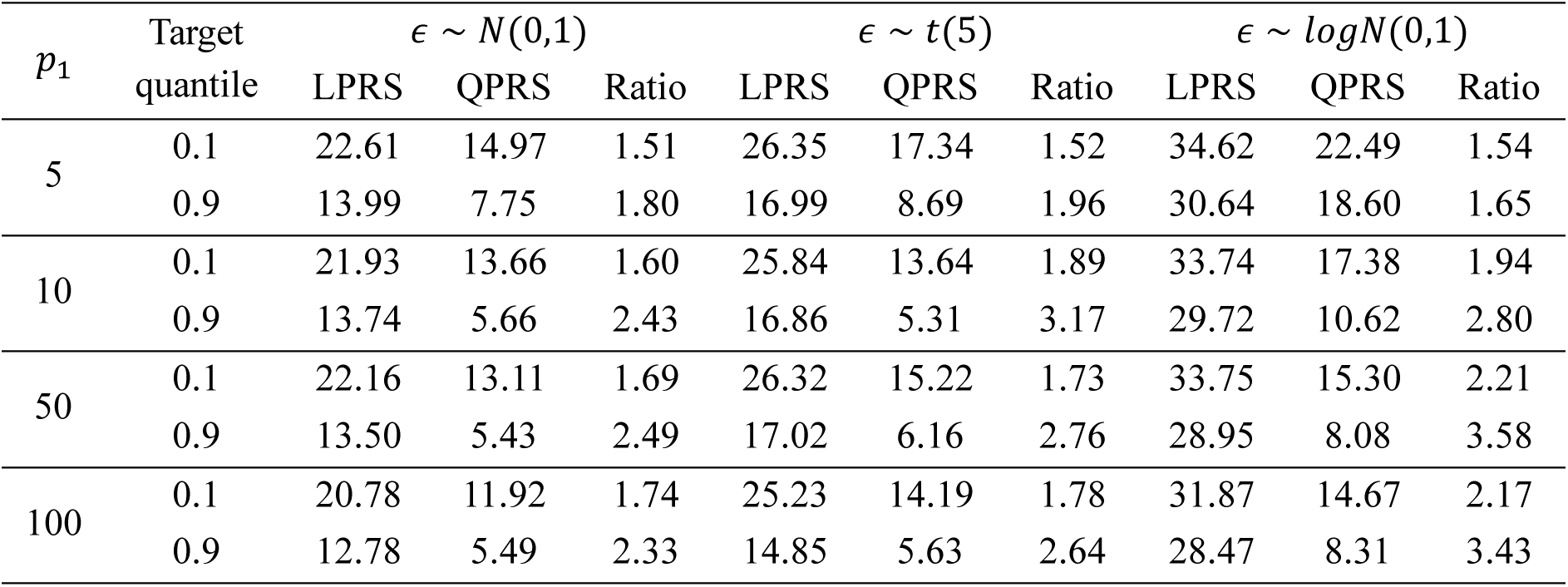
Mean of MSE of LPRS and QPRS in simulation Scheme 1 across varying numbers of causal SNPs ( *p*_1_) and error distributions ( *ε* ∼ *N*(0,1), *t*(5), *logN*(0,1)). The ratio indicates relative performance (LPRS/QPRS).

#### Scheme 2: With Outliers

Table 3 reports the TPR, FPR, and MSE across varying proportions and magnitudes of outlier effects. The results highlight a trade-off between sensitivity and specificity that differs across methods and levels of contamination.

**Table 3.** TPR, FPR, and MSE for mean regression and median regression in Scheme 2 across varying outlier proportions and magnitudes. FPR values are multiplied by 10^2^.

|  | Outlier Degree | 5 |  |  |  | 10 |  |  |  | 20 |  |  |  |
| --- | --- | --- | --- | --- | --- | --- | --- | --- | --- | --- | --- | --- | --- |
| Outlier % | Method | TPR | FPR | MSE (mean) | MSE (med) | TPR | FPR | MSE (mean) | MSE (med) | TPR | FPR | MSE (mean) | MSE (med) |
| 1 | LPRS | 0.474 | 0.162 | 23.65 | 24.46 | 0.360 | 0.016 | 23.13 | 24.82 | 0.194 | 0.007 | 22.78 | 26.54 |
|  | QPRS | 0.382 | 0.015 | 19.97 | 20.55 | 0.382 | 0.015 | 19.77 | 21.38 | 0.382 | 0.015 | 20.23 | 23.41 |
| 2 | LPRS | 0.360 | 0.021 | 23.64 | 24.76 | 0.294 | 0.010 | 23.37 | 26.45 | 0.140 | 0.004 | 23.57 | 30.22 |
|  | QPRS | 0.372 | 0.015 | 19.80 | 20.96 | 0.424 | 0.021 | 20.36 | 22.85 | 0.376 | 0.015 | 22.26 | 27.32 |
| 5 | LPRS | 0.194 | 0.023 | 23.84 | 26.63 | 0.224 | 0.006 | 23.59 | 31.64 | 0.118 | 0.002 | 25.57 | 45.53 |
|  | QPRS | 0.508 | 0.064 | 20.30 | 22.78 | 0.418 | 0.021 | 22.14 | 28.68 | 0.510 | 0.066 | 28.10 | 44.03 |
| 10 | LPRS | 0.402 | 0.019 | 23.50 | 30.03 | 0.216 | 0.005 | 24.46 | 43.56 | 0.132 | 0.002 | 25.86 | 85.39 |
|  | QPRS | 0.362 | 0.016 | 21.50 | 27.12 | 0.512 | 0.101 | 25.20 | 41.38 | 0.360 | 0.052 | 33.56 | 83.85 |

Under mild contamination (1–2 of observations), LPRS initially appears more sensitive, achieving a higher TPR than QPRS, particularly when the outlier magnitude is smallest (i.e., a multiplication factor of 5). However, this gain in sensitivity comes at the cost of a substantially inflated FPR (up to 0.162), indicating a high rate of false discoveries. In contrast, QPRS adopts a more conservative approach, maintaining a consistently low FPR. As contamination becomes more severe (5–10 of observations), the advantage of QPRS’s robustness becomes evident. The performance of LPRS degrades sharply, with its TPR collapsing to as low as 0.118. Conversely, QPRS demonstrates resilience, often achieving a substantially higher TPR than LPRS in these settings while still maintaining a lower FPR. Overall, these findings indicate that while LPRS is more sensitive under light contamination, it is also more prone to false positives. In contrast, QPRS maintains lower FPRs and demonstrates greater resilience in detecting true signals as outlier severity increases.

It is worth noting that the variant selection patterns observed in scheme 2 (Table 3, TPR and FPR) differ substantially from those in scheme 1 (Table 1). This discrepancy arises from differences in the underlying data-generating mechanisms. In scheme 1, SNPs contribute to vQTLs, creating quantile-dependent signals that are naturally captured by QPRS but largely missed by LPRS, which is restricted to mean effects. As a result, QPRS identifies causal variants more effectively, even at the expense of slightly elevated FPR (see Table 1). In contrast, scheme 2 is generated under a homogeneous mean-effect model, *β*(*τ*) = *β* for all *τ* ∈ (0,1), where true SNP effects do not vary across quantiles. In this setting, QPRS does not have an advantage in capturing signal heterogeneity, but its robustness to outliers helps preserve predictive accuracy (Table 3). Under light contamination, LPRS can sometimes recover a higher proportion of causal SNPs, albeit at the cost of increased false positives. QPRS, in contrast, tends to trade sensitivity for specificity, maintaining lower FPR while achieving moderate TPR. As contamination becomes more severe, the stability of QPRS becomes more apparent, with TPR often surpassing that of LPRS, as the latter’s performance declines sharply under heavy outlier influence.

For predictive performance, QPRS achieves lower MSE than LPRS in both mean and median prediction across almost all settings. When outlier contamination is mild (1–2 of observations), QPRS shows a clear advantage, reducing MSE compared with LPRS. At moderate contamination levels (5 of observations), the improvement remains evident, with ratios around 1.10–1.17 in median regression depending on outlier magnitude. Under more extreme contamination (10 of observations), the difference narrows; ratios approach 1.02–1.11 in median regression, indicating that both methods degrade under heavy contamination, though QPRS still maintains a modest advantage. A similar pattern is observed in the mean prediction task.

Overall, the comparison of schemes 1 and 2 illustrates that QPRS is particularly advantageous when genetic effects vary across quantiles, as in variance QTL (vQTL) settings. In contrast, under homogeneous mean-effect models subject to outlier contamination, its main strength lies in robustness to contamination and in preserving specificity, rather than in consistently achieving higher sensitivity.

### 3.2 Real Data Analysis Results

The phenotype we examined is 2-hour post-OGTT blood glucose (mg/dL). The distribution of glucose levels is right-skewed: most observations are below 200 mg/dL, with a small number of markedly high values. This skewness limits the suitability of linear regression for modeling this trait. Although a log transformation could be applied to approximate normality, it complicates effect-size interpretation and does not adequately capture heterogeneity across the distribution. Moreover, despite applying a logarithmic transformation to the phenotype, the Kolmogorov–Smirnov test indicates that the distribution still deviates significantly from normality. Our proposed method achieves robust predictive performance because QR does not rely on strong distributional assumptions.

We first present the SNPs that QR-GWAS identifies. Figure 2 (top panel) displays overlaid Manhattan plots from QR-GWAS at *τ* = 0.1, 0.2, …, 0.9. Compared with conventional linear regression GWAS (bottom panel), QR-GWAS detects a broader set of significant variants, including many that linear regression fails to identify. While individual quantiles may yield fewer signals than linear regression-based GWAS, combining results across quantiles reveals variants whose effects are obscured by heterogeneity or non-normality.

**Figure 2.**
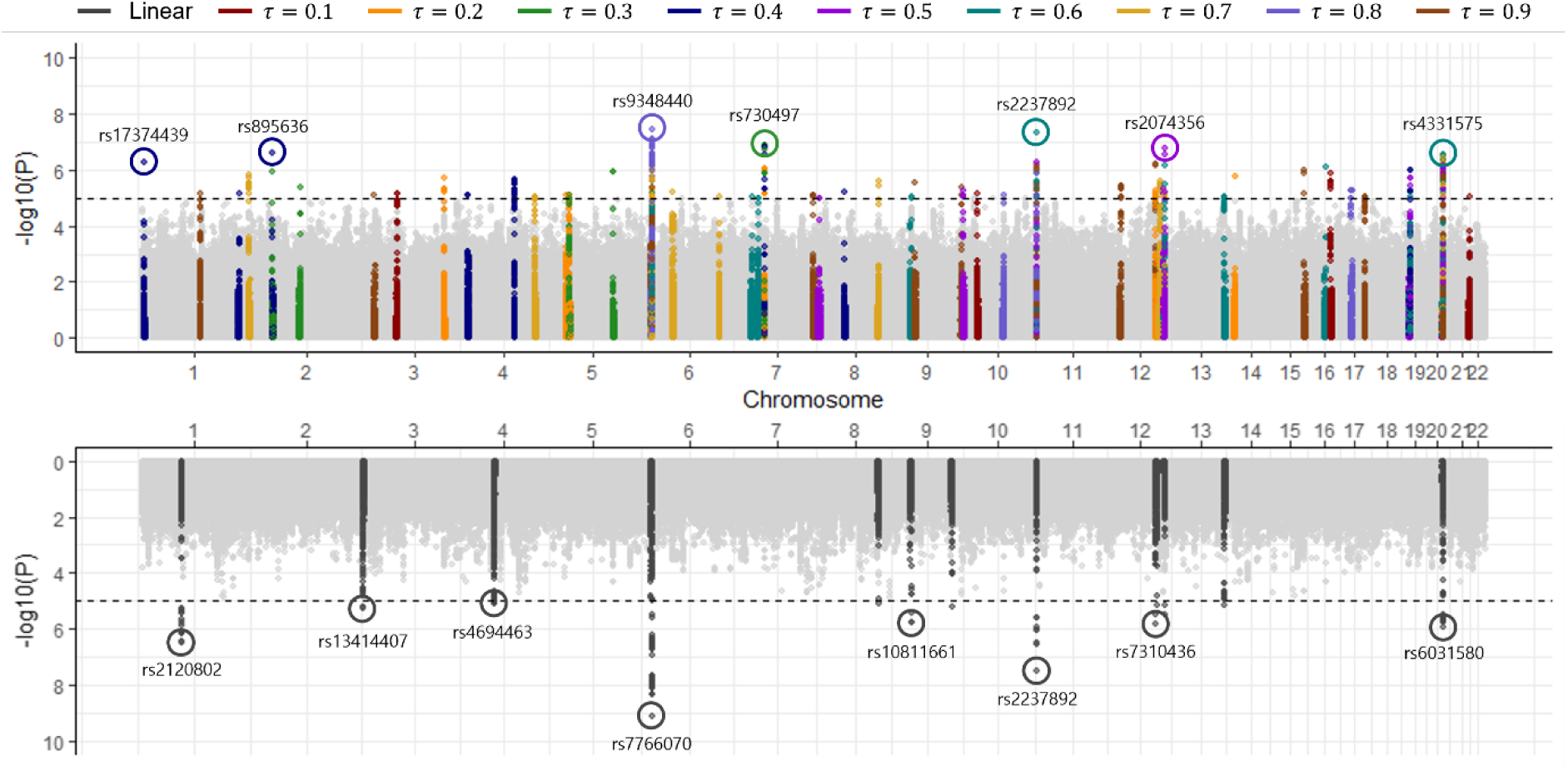
Manhattan plots of GWAS for 2-hour post-OGTT glucose levels. The top panel shows overlaid *p*-values from QR-GWAS across nine quantiles. The bottom panel shows association results from a standard linear regression-based GWAS.

Figure 3 presents an UpSet plot illustrating overlaps among significant SNPs detected by LPRS and QPRS. The horizontal bars display the total number of SNPs identified by each method, while vertical bars show intersections across methods. In the dot matrix below, filled circles indicate the sets included in each intersection, with connecting lines denoting the specific combinations compared. QPRS identifies 342 significant SNPs, of which 77 overlap with LPRS, while 73 are unique to LPRS and 265 are unique to QPRS. Notably, 56 SNPs are uniquely identified at QPRS(0.4), while additional unique discoveries also appear at other quantiles, particularly QPRS(0.1) (23 SNPs) and QPRS(0.9) (27 SNPs). The concentration of discoveries at *τ* = 0.4 is likely attributable to the right-skewed nature of the 2-hour glucose distribution in our data. In such a distribution, the mode, the point of highest data density, is typically located to the left of the median (*τ* = 0.5). Consequently, the τ=0.4 quantile falls within this high-density region. Although tail quantiles tend to have lower power, they can reveal distribution-specific effects. The discoveries at QPRS(0.1) and QPRS(0.9) suggest that certain genetic effects are more apparent in the lower or upper tails of the phenotype distribution.

**Figure 3.**
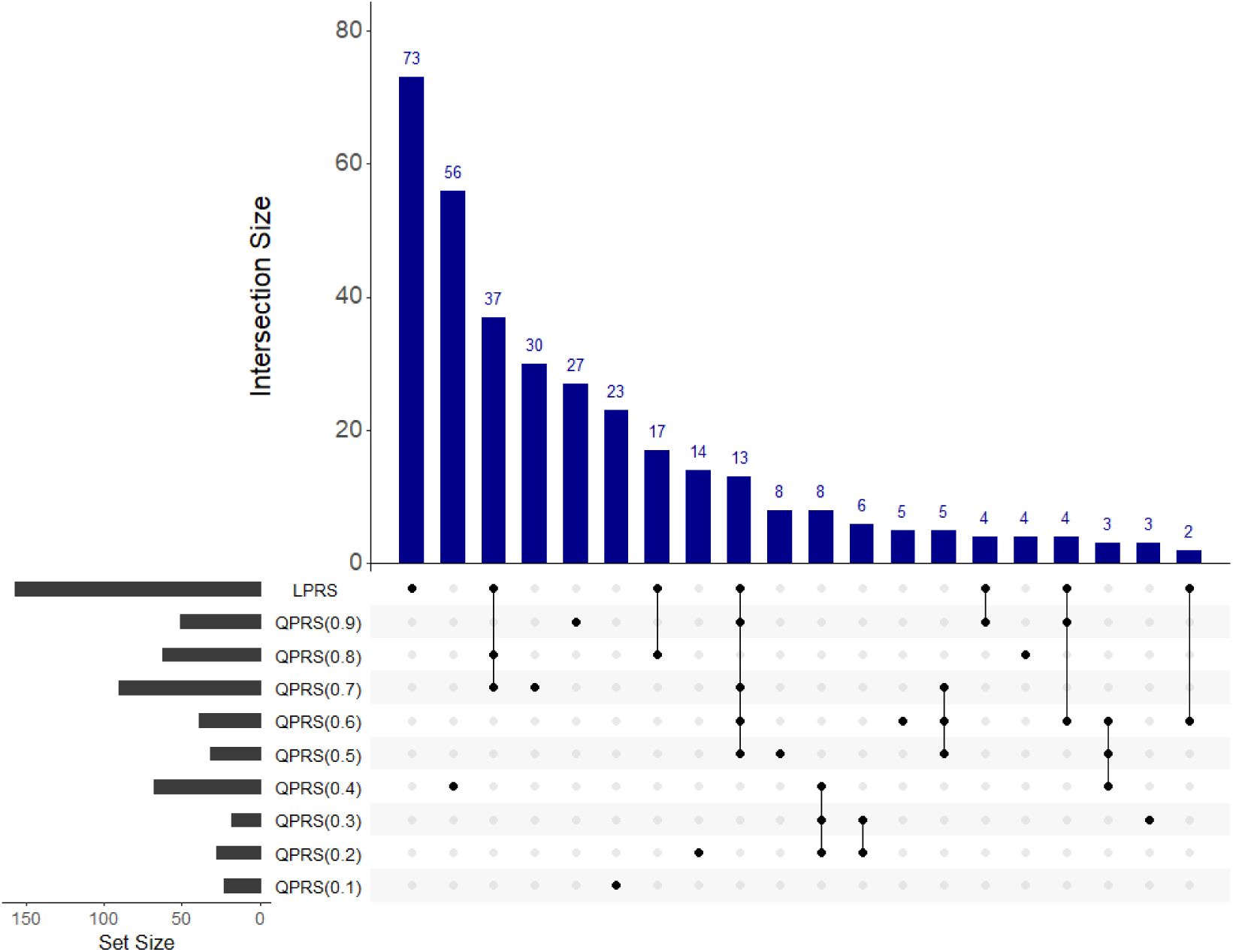
Upset plot of SNP overlaps among LPRS and QPRS from τ = 0.1 to 0.9. The horizontal bars (set size) show the total number of SNPs with p-values below 1 × 10^−5^ identified by each PRS. In the dot matrix below, filled circles indicate the PRSs involved in each intersection, and the vertical bars (intersection size) represent the number of SNPs shared across those models.

Two examples of SNPs with heterogeneous effects are rs7306855 and rs17153083. Their effect sizes vary dramatically across the phenotype distribution in a pattern completely obscured by the conventional mean effect. For SNP rs7306855, the effect is strongly positive in the lower quantiles (*β̂*= 6.72 at *τ* = 0.1), suggesting it raises glucose levels for individuals at the lower end of the distribution. However, this effect diminishes and sharply reverses, becoming strongly negative in the upper tail (*β̂*= −15.00 at *τ* = 0.9). Similarly, SNP rs17153083 exhibits a strongly negative effect at the highest quantile (*β̂* = −10.22 at *τ* = 0.9).

In both cases, the conventional mean regression provides a single, uninformative estimate (*β̂* = −0.44 and 0.05, respectively) that fails to capture these complex relationships. These patterns are representative examples of heterogeneous effects. The signals from these SNPs are largely obscured in a conventional mean regression but are clearly revealed when modeling quantile-specific effects. These examples, therefore, illustrate the type of complex genetic signal that motivates the QPRS framework; they highlight that QPRS can capture heterogeneous genetic influences that conventional PRS methods are not designed to detect.

We next assess the predictive performance of QPRS relative to four alternatives: LPRS, Lassosum, LDpred, and SBLUP. As outlined in Section 2, performance is evaluated under two frameworks: (a) PRS-only models and (b) models adjusted for covariates including sex, age, BMI, smoking status, and the first 10 principal components. Table 4 reports the results from 5-fold cross-validation. For the continuous trait, 2-hour post-OGTT glucose, QPRS attains consistently higher *R*^2^ than competing methods across both frameworks. The combined model, which includes both LPRS and QPRS, yields the strongest predictive performance. This finding suggests that QPRS provides complementary information to LPRS. A similar pattern is observed for the binary trait. QPRS and the combined model achieve superior accuracy compared with other approaches across both the observed-scale AUC and the more robust liability-scale AUC (AUCL). For instance, in the covariate-adjusted models, QPRS attains the highest AUC (0.6675) and AUCL (0.5634) among all individual methods. Again, the combined model performs best, reaching an AUC of 0.6838 and an AUCL of 0.5699. Overall, these results demonstrate that QPRS can enhance polygenic prediction by incorporating distributional information beyond mean effects.

**Table 4.**
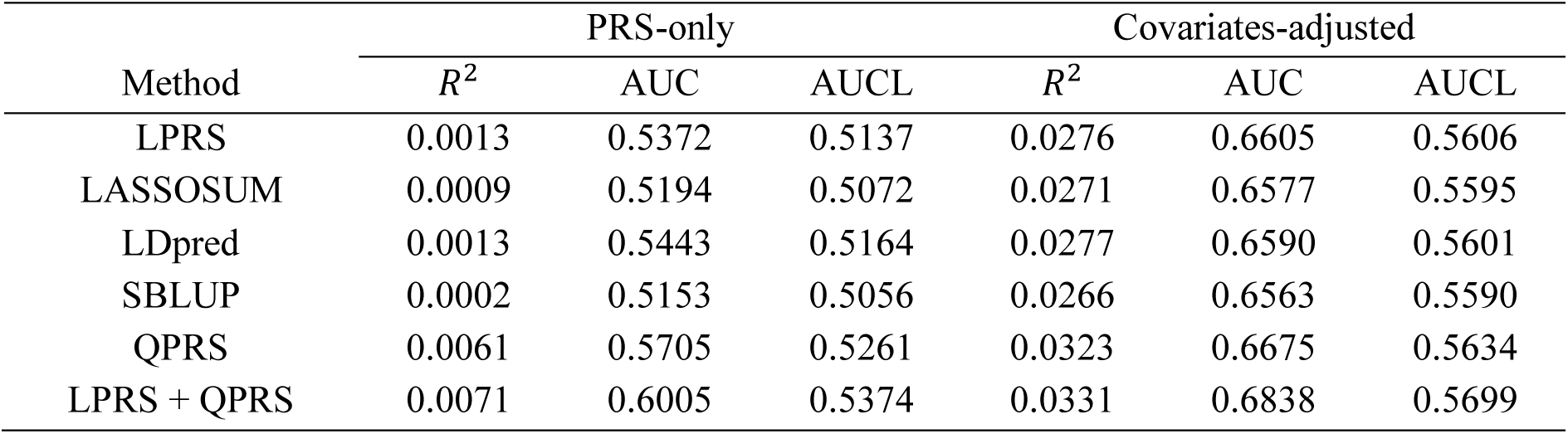
Predictive Performance of polygenic risk score methods under PRS-only and covariate-adjusted (with sex, age, BMI, smoking status, 10 PCs) models. AUCL denotes liability AUC.

To investigate whether QPRS provides unique predictive information not captured by LPRS, we evaluate predictive accuracy by sequentially adding QPRS components to a baseline LPRS-only model. Figure 4 shows the incremental gains from adding each QPRS(*τ*) from *τ* = 0.1 to 0.9. While the contributions of lower and middle quantiles vary, their cumulative addition consistently enhances prediction compared with LPRS alone. Notably, QPRS(0.9) provides the largest improvement. In the covariate-adjusted model for the continuous trait, adding QPRS(0.9) increases *R*^2^ by 0.0018. For the binary trait, adding QPRS(0.9) raises AUC by 0.0079 in the covariate-adjusted model and by 0.00178 in the PRS-only setting These gains reflect the right-skewed distribution of the phenotype, where linear regression underperforms due to data sparsity in the upper tail. In contrast, QPRS is better able to capture genetic effects in such regions. These results align with recent studies applying QR to investigate genetic effects at distributional extremes^17,19,35^. Overall, our findings underscore the value of quantile-specific risk scores for improving phenotype prediction, particularly in skewed traits such as the 2-hour post-OGTT glucose levels analyzed here, where clinical risk is concentrated in the upper tail of the distribution.

**Figure 4.**
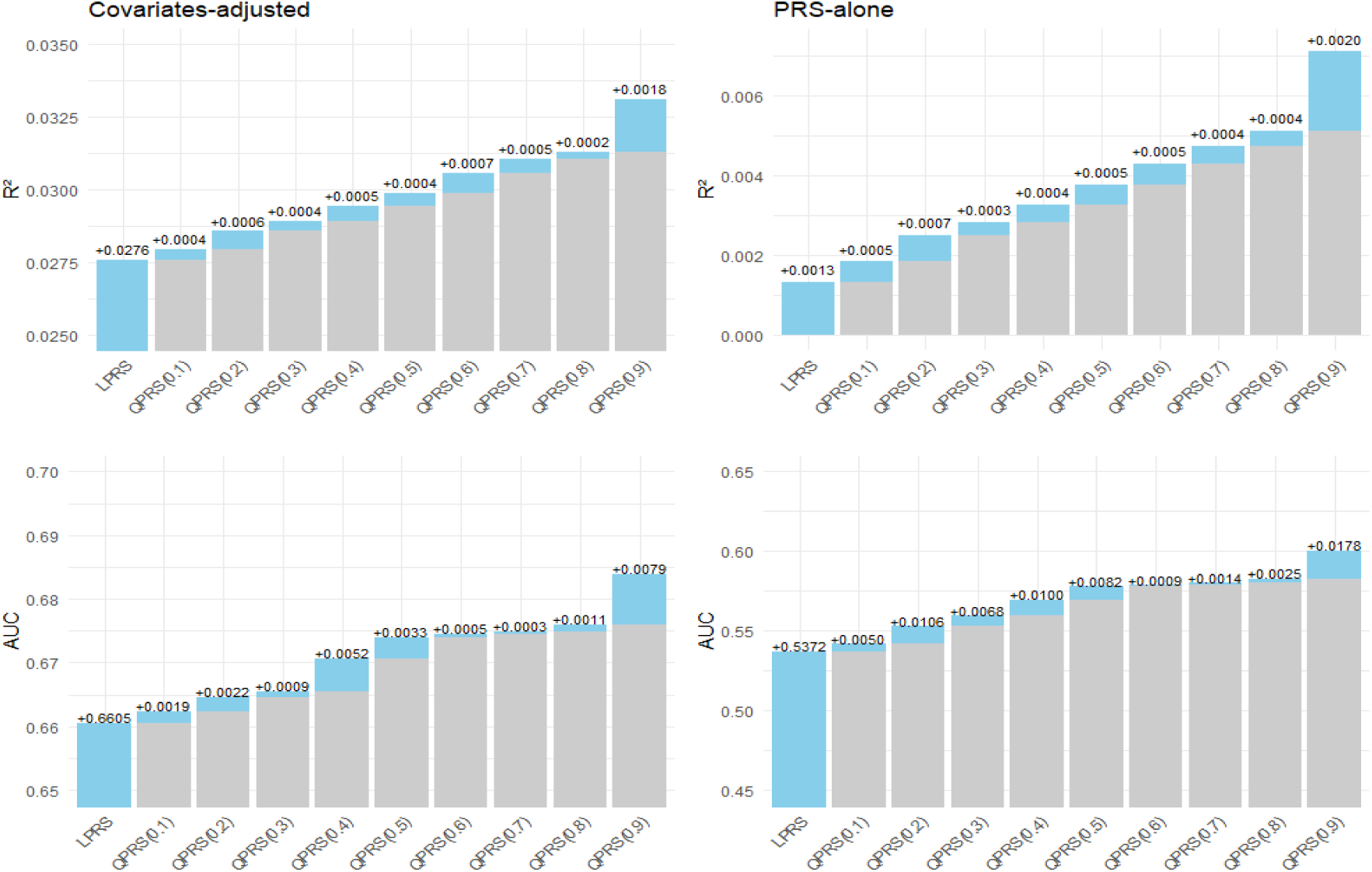
Incremental predictive performance by sequentially adding QPRS components. Top/bottom rows show *R*^2^ (continuous trait) and AUC (binary trait), and left/right columns show covariate-adjusted and PRS-only models, respectively. QPRS(*τ*) at *τ* = 0.1, 0.2, …,0.9 are sequentially added to a baseline LPRS model. Values above bars indicate the incremental contribution of each newly added QPRS(*τ*).

## 4. Discussion

This study highlights the importance of modeling heterogeneity in genetic effects when constructing PRSs. By incorporating QR into the PRS framework, we show that genetic influences can vary systematically across the phenotype distribution, with implications for both prediction and discovery. The observed gains in predictive accuracy, particularly in the presence of skewed phenotypes such as 2-hour post-OGTT glucose, illustrate that mean-based models may underestimate genetic contributions in distributional extremes.

The ability of QR-GWAS to identify variants missed by linear models further emphasizes the biological relevance of distribution-specific effects. These findings align with emerging evidence that genetic associations may manifest through interactions with phenotypic scale, environment, or unmeasured modifiers, resulting in non-uniform effects across quantiles. In this light, QPRS does not merely refine prediction, but also provides an additional lens for exploring the architecture of complex traits.

From a methodological standpoint, our approach complements existing PRS strategies rather than replacing them. The consistent improvement when QPRS is combined with conventional PRS suggests that distinct layers of information are captured by mean- and quantile-based modeling. This complementarity may prove particularly valuable in precision medicine, where capturing variability across risk strata is crucial for effective stratification.

Several extensions naturally follow. Penalized^36^ or Bayesian^37^ QR could be incorporated to improve SNP selection beyond clumping and thresholding. Moreover, validation across multiple traits and ancestries will be necessary to establish the generalizability of QPRS. Finally, as biobank-scale data continue to expand, QPRS provides a scalable framework for integrating distributional heterogeneity into large-scale genetic prediction. Developing a Lassosum-like approach to leverage large-scale QR-GWAS summary statistics is an important direction for future work.

## Acknowledgements

This study was conducted with bioresources from National Biobank of Korea, the Korea Disease Control and Prevention Agency, Republic of Korea (NBK-2021-059).

## Author Contributions

Conceptualization: MP. Methodology: MP, SK. Software and simulations: SK. Data curation and formal analysis: TG. Funding acquisition: MP. Writing - original draft: SK, MP. Writing - review & editing: TP, MP.

## Data Availability Statement

Genotype data from the Korea Association Resource (KARE) cohort, which is part of the Korean Genome and Epidemiology Study (KoGES), were accessed through the Clinical and Omics Data Archive (CODA; https://coda.nih.go.kr) of the Korea Disease Control and Prevention Agency (KDCA).

## References

1 Choi, S. W., Mak, T. S.-H. & O’Reilly, P. F. Tutorial: a guide to performing polygenic risk score analyses. Nature protocols 15, 2759–2772 (2020).

2 Cross, B., Turner, R. & Pirmohamed, M. Polygenic risk scores: An overview from bench to bedside for personalised medicine. Frontiers in genetics 13, 1000667 (2022).

3 Jia, G., et al. Evaluating the utility of polygenic risk scores in identifying high-risk individuals for eight common cancers. JNCI cancer spectrum 4, pkaa021 (2020).

4 Mak, T. S. H., Porsch, R. M., Choi, S. W., Zhou, X. & Sham, P. C. Polygenic scores via penalized regression on summary statistics. Genetic epidemiology 41, 469–480 (2017).

5 Vilhjálmsson, B. J. et al. Modeling linkage disequilibrium increases accuracy of polygenic risk scores. The american journal of human genetics 97, 576–592 (2015).

6 Robinson, M. R. et al. Genetic evidence of assortative mating in humans. Nature Human Behaviour 1, 0016 (2017).

7 Wang, Z. et al. The value of rare genetic variation in the prediction of common obesity in European ancestry populations. Frontiers in endocrinology 13, 863893 (2022).

8 Zhang, Q., Privé, F., Vilhjálmsson, B. & Speed, D. Improved genetic prediction of complex traits from individual-level data or summary statistics. Nature communications 12, 4192 (2021).

9 Albert, E. et al. Transferability of the PRS estimates for height and BMI obtained from the European ethnic groups to the Western Russian populations. Frontiers in genetics 14, 1086709 (2023).

10 Ren, J., Lin, Z., He, R., Shen, X. & Pan, W. Using GWAS summary data to impute traits for genotyped individuals. Human Genetics and Genomics Advances 4 (2023).

11 Miao, J. et al. A quantile integral linear model to quantify genetic effects on phenotypic variability. Proceedings of the National Academy of Sciences 119, e2212959119 (2022).

12 Shi, G. Genome-wide variance quantitative trait locus analysis suggests small interaction effects in blood pressure traits. Scientific Reports 12, 12649 (2022).

13 Westerman, K. E. et al. Variance-quantitative trait loci enable systematic discovery of gene-environment interactions for cardiometabolic serum biomarkers. Nature communications 13, 3993 (2022).

14 Yang, J. et al. FTO genotype is associated with phenotypic variability of body mass index. Nature 490, 267–272 (2012).

15 Koenker, R. & Bassett Jr, G. Regression quantiles. Econometrica: journal of the Econometric Society, 33–50 (1978).

16 Koenker, R. & Hallock, K. F. Quantile regression. Journal of economic perspectives 15, 143–156 (2001).

17 Nascimento, M. et al. Quantile regression for genome-wide association study of flowering time-related traits in common bean. PLoS One 13, e0190303 (2018).

18 Wang, F. et al. Regenie. QRS: computationally efficient whole-genome quantile regression at biobank scale. bioRxiv, 2025.2005. 2002.651730 (2025).

19 Wang, C. et al. Genome-wide discovery for biomarkers using quantile regression at biobank scale. Nature Communications 15, 6460 (2024).

20 Mefford, J. et al. Beyond predictive R2: Quantile regression and non-equivalence tests reveal complex relationships of traits and polygenic scores. The American Journal of Human Genetics 112, 1363–1375 (2025).

21 Consortium, G. P. A global reference for human genetic variation. Nature 526, 68 (2015).

22 Purcell, S. M. et al. Common polygenic variation contributes to risk of schizophrenia and bipolar disorder. Nature 460, 748–752, doi:10.1038/nature08185 (2009).

23 Dudbridge, F. Power and predictive accuracy of polygenic risk scores. PLoS genetics 9, e1003348 (2013).

24 Koenker, R. & Bassett Jr, G. Robust tests for heteroscedasticity based on regression quantiles. Econometrica: Journal of the Econometric Society, 43–61 (1982).

25 Koenker, R. Quantile regression. Vol. 38 (Cambridge university press, 2005).

26 Wray, N. R., Goddard, M. E. & Visscher, P. M. Prediction of individual genetic risk to disease from genome-wide association studies. Genome research 17, 1520–1528 (2007).

27 Purcell, S. et al. PLINK: a tool set for whole-genome association and population-based linkage analyses. The American journal of human genetics 81, 559–575 (2007).

28 Choi, S. W. & O’Reilly, P. F. PRSice-2: Polygenic Risk Score software for biobank-scale data. Gigascience 8, giz082 (2019).

29 Committee, A. D. A. P. P. 2. Diagnosis and Classification of Diabetes: Standards of Care in Diabetes—2025. Diabetes Care 48, S27–S49, doi:10.2337/dc25-S002 (2024).

30 Menke, A., Rust, K. F., Savage, P. J. & Cowie, C. C. Hemoglobin A1c, fasting plasma glucose, and 2-hour plasma glucose distributions in US population subgroups: NHANES 2005–2010. Annals of epidemiology 24, 83–89 (2014).

31 Lee, S. H., Goddard, M. E., Wray, N. R. & Visscher, P. M. A better coefficient of determination for genetic profile analysis. Genetic epidemiology 36, 214–224 (2012).

32 Koenker, R., et al. Package ‘quantreg’. Reference manual available at R-CRAN: https://cran.rproject.org/web/packages/quantreg/quantreg.pdf (2018).

33 Yang, J., Lee, S. H., Goddard, M. E. & Visscher, P. M. GCTA: a tool for genome-wide complex trait analysis. The American Journal of Human Genetics 88, 76–82 (2011).

34 Loh, P.-R. et al. Efficient Bayesian mixed-model analysis increases association power in large cohorts. Nature genetics 47, 284–290 (2015).

35 Pozarickij, A., Williams, C., Hysi, P. G. & Guggenheim, J. A. Quantile regression analysis reveals widespread evidence for gene-environment or gene-gene interactions in myopia development. Communications biology 2, 167 (2019).

36 Belloni, A. & Chernozhukov, V. ℓ 1-penalized quantile regression in high-dimensional sparse models. (2011).

37 Yu, K. & Moyeed, R. A. Bayesian quantile regression. Statistics & Probability Letters 54, 437–447 (2001).

